# Zoonotic disease preparedness among frontline healthcare workers in Ghana: Assessing literacy and behaviors in a One Health context

**DOI:** 10.1101/2025.07.08.25331119

**Authors:** Kirstin P. West, Kristin K. Sznajder, Hannah E. Sauve, Grace Hwang, Kedir Teji Roba, Leonard Baatiema, Ernest Kenu, Charles L. Noora, Abraham Tamirat Gizaw, Abebayehu N. Yilma

**Author notes:** **Correspondence:** Abebayehu N. Yilma, PhD MPH.

## Abstract

Zoonotic diseases pose a growing global health threat, particularly in low-resource settings where human-animal-environmental interfaces are common. Although Ghana has adopted One Health policies, limited evidence exists on frontline healthcare workers’ (FHWs) zoonosis preparedness. Thus, this study assessed FHWs’ knowledge, attitudes, and practices related to zoonoses and One Health in Ghana’s Ada East district. We conducted a cross-sectional survey of 250 FHWs across 18 selected health facilities. A 54-item questionnaire measured knowledge, attitudes, and practices regarding priority zoonoses, Ebola virus disease, and the One Health approach. Scores were scaled to 100% and dichotomized using Bloom’s cut-off, with >60% representing “good knowledge,” “positive attitudes,” or “good practices.” Multivariable logistic regression identified predictors of good knowledge and practices. According to this study, while 79.2% of FHWs scored ≥60% on the total knowledge scale, classified as “good knowledge” of zoonoses, Ebola virus disease, and One Health, only 22.4% correctly identified >60% of listed zoonoses. Ebola-specific knowledge was high, though misconceptions about transmission and treatment persisted. Only 6.8% scored ≥60% on the attitude scale, reflecting limited confidence and widespread concern about personal safety, institutional readiness, and community stigma. While 47.2% demonstrated “good” prevention and control practices, institutional protocol use and intersectoral collaboration were limited. Rabies-focused training increased odds of good knowledge (AOR = 3.25, 95% CI: 1.04–11.1) and practices (AOR = 2.46, 95% CI: 1.07–5.87). In-facility training improved practices (AOR = 2.77, 95% CI: 1.34–5.92), whereas government-delivered training and infrequent climate-health engagement were linked to poor knowledge and practices. Gaps in zoonoses literacy and institutional preparedness were evident among FHWs. Targeted staff retraining, peer-led mentorship, and expanded rabies training may offer practical entry points for One Health capacity building. Addressing stigma and embedding climate-health content into community-based risk communication may improve frontline readiness.

## Introduction

Zoonotic diseases, infections transmissible between animals and humans, present a major and growing global health threat. Nearly 60% of all known human pathogens and around three-quarters of new or emerging infectious diseases have animal origins (1–5). Each year, zoonoses contribute to nearly 2.5 billion illnesses and 3 million deaths worldwide, spanning both endemic threats such as rabies and pandemic events like COVID-19 (2,6,7). An estimated 98% of the global zoonotic disease burden is shouldered by developing regions, where zoonoses account for more than ten million disability-adjusted life-years (DALYs) annually (2,6,8). Limited public health infrastructure and frequent human-animal interactions continue to elevate the risk of zoonotic spillover in these regions (9–13).

Sub-Saharan Africa (SSA) experiences a particularly high zoonotic disease burden, with a systematic review of outbreaks from 2000 to 2022 reporting a pooled infection rate of 54 per 1,000 people and a case fatality rate of 345 per 1,000 infected individuals (2,6,14,15). Between 2012 and 2022, the continent recorded a 63% increase in zoonotic outbreaks compared to the previous decade (16). The West African Ebola virus disease (EVD) epidemic of 2014 — 2016, with over 28,600 cases and 11,300 deaths across Guinea, Liberia, and Sierra Leone, illustrated how rapidly zoonotic spillovers can evolve into widespread global health crises (17–20).

Ghana, located in West Africa, remains highly vulnerable to zoonotic spillovers. Notable endemic zoonoses in Ghana include rabies, influenza, salmonellosis, anthrax, brucellosis, and bovine tuberculosis, while viral hemorrhagic fevers such as Ebola and Lassa fever pose persistent risks (15,21). The 2022 confirmation of Marburg virus in Ghana marked the country’s first recorded outbreak of a filovirus pathogen, highlighting the region as a potential hotspot for zoonotic diseases (22,23).

This heightened risk of zoonoses is shaped by frequent and culturally embedded human-animal interactions. Common practices such as livestock rearing, bushmeat hunting, and habitat encroachment bring people into close contact with wildlife reservoirs (24,25). Frugivorous bat species, natural hosts of EVD and Marburg viruses, are prevalent in Ghana (25,26). A 2011 – 2012 survey found that about 47% of respondents visit bat caves and about 46% consume bat meat in the region (27,28). Nearly half of Ghanaians engage in agriculture or animal husbandry (29), and bushmeat trade remains culturally ingrained (30–33), especially in the Greater Accra Region, where peri-urban farming, livestock markets, and transport corridors facilitate cross-species transmission (25).

Recognizing the multifaceted drivers of zoonotic emergence, Ghana adopted a One Health approach in 2018 as a strategic framework for improving health security and promoting cross-sector collaboration among health, veterinary, wildlife, and environmental sectors to jointly address zoonotic threats (34–37). Specifically, Ghana has established a national One Health platform, coordinated surveillance, and engaged in the Global Health Security Agenda to strengthen capacities for zoonotic disease prevention and response (34,36,38,39). Yet, knowledge and preparedness gaps regarding zoonotic diseases persist among frontline healthcare workers (FHWs). For instance, many remain hesitant to manage EVD cases because of fear and limited training (36,40–42). While the One Health approach has been adopted at the policy level in Ghana, translating this national framework into effective frontline knowledge, attitudes, and practices (KAP) remains an ongoing challenge, and further efforts are needed to ensure effective integration (15,25,36,37,42–44).

FHWs play a critical role in the early detection, reporting, and control of zoonotic disease outbreaks (45). As the first point of contact for patients, FHWs’ KAP regarding priority and emerging zoonoses can greatly influence their ability to identify zoonotic cases, implement infection prevention and control measures, and alert public health authorities (45). However, recent data from Northern Ghana suggests that while about 80% of healthcare workers knew of zoonotic diseases, only 18.7% could identify multiple zoonotic diseases, and 59% understood One Health principles (41). Although previous studies have explored zoonotic disease and One Health knowledge among healthcare workers, these analyses primarily assessed knowledge levels and considered professional experience as a predictor of knowledge (40,41).

This formative baseline study contributes to existing literature by assessing and characterizing not only knowledge but also attitudes and practices of FHWs related to zoonotic diseases and One Health within the Greater Accra Region —Ghana’s capital— where active human-animal interactions increase the risk of zoonotic transmission. We examine additional factors, specifically formal training and access to health-related information, as predictors of KAP performance. Findings will inform targeted training needs, risk communication strategies, and frontline-level One Health policy implementation, thereby addressing preparedness gaps and strengthening Ghana’s readiness in an era of ever-emerging zoonotic threats.

## Materials and methods

### Study Design and Setting

This study used a cross-sectional survey to assess the KAP of FHWs regarding zoonotic diseases, with a focus on the One Health approach and EVD. Data were collected between January 30^th^, 2025, and February 7^th^, 2025, in Ghana’s Greater Accra Region, specifically within the Ada East district. This region includes the nation’s capital, which comprises densely populated urban centers, peri-urban and rural communities (46). Ada East was selected for its high concentration of healthcare facilities and diverse healthcare workforce, including nurses, midwives, pharmacists, community health officers, physicians, among many other cadres of clinical staff. In collaboration with the University of Ghana, eighteen facilities —including district-level hospitals, local health centers, and community-based health planning and services (CHPS) compounds— were purposively sampled to capture this diversity (Figure 1). Several of these sites routinely manage infectious diseases and serve as key points for outbreak detection in the country.

**Fig 1.**
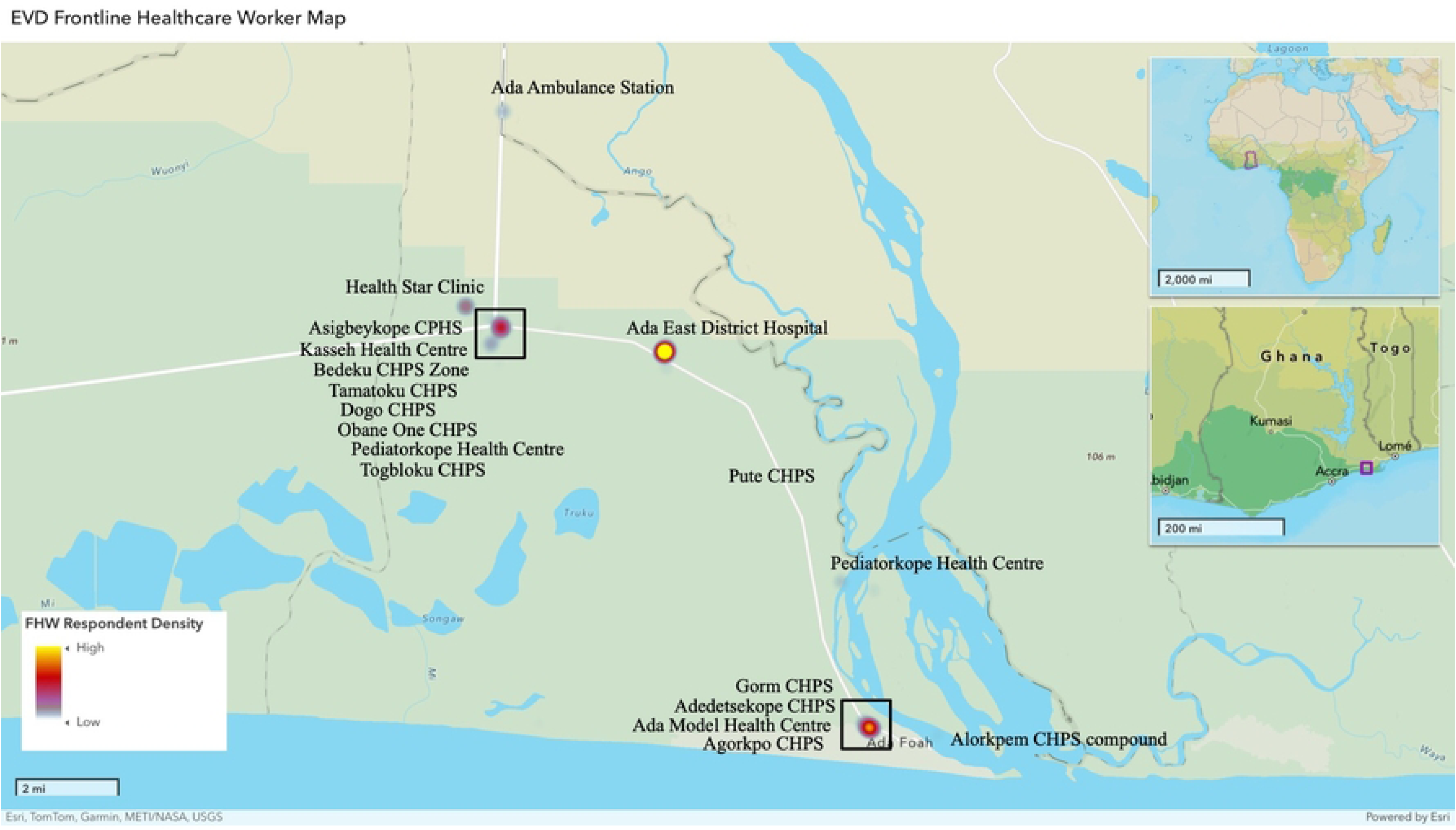
Geographic Distribution of Surveyed Health Facilities and Frontline Healthcare Worker Respondents in Ada East District, Ghana. This map displays the 18 health facilities included in the study —comprising district hospitals, local health centers, and CHPS compounds— across the Ada East district. Facilities were purposively sampled to reflect the range of care settings in the district. Locations were geocoded and mapped using ArcGIS Online v3.2. Color gradients indicate the density of participating frontline healthcare workers (FHWs), with yellow representing high respondent density, red medium, and blue low. Facility names are labeled to illustrate spatial coverage.

### Study Population

All eligible FHWs at the selected healthcare facilities were invited to participate. The study population included FHWs actively employed at participating sites in the Greater Accra Region. For this study, FHWs were defined as clinical personnel directly involved in patient care and likely to serve as first responders during infectious disease outbreaks. This encompassed clinical staff (e.g., physicians, nurses, midwives, physician assistants), clinical support staff (e.g., laboratory technologists, biomedical scientists), and other patient-facing cadres (e.g., pharmacists, radiographers, emergency response teams).

Participants were eligible if they: (1) were ≥18 years old; (2) held a frontline clinical role at a selected facility; (3) had ≥3 months of experience at their current site; (4) were ≥6 months post-training; (5) could read, write, and speak English; and (6) provided written informed consent. Individuals not directly involved in patient care, unavailable during data collection, or unwilling to participate were excluded.

### Sample Size Calculation

Due to the lack of prior studies in the study area, the required sample size was estimated using a single population proportion formula to ensure adequate power for estimating KAP assessments. Assuming 6% of the population had poor KAP toward zoonotic disease and One Health, with a 3% margin of error and 95% confidence interval (CI), the minimum required sample size was 241. To account for potential nonresponse and ensure adequate representation, a target sample size of 250 participants was set.

### Participant Recruitment and Sampling

Recruitment was conducted on-site at each selected healthcare facility. Before data collection, the research team coordinated with facility managers and department heads to identify optimal times for recruitment based on clinic workflows and shift schedules. Trained local research assistants (RAs) were stationed in staff common areas during both day and night shifts, distributing information sheets and educational leaflets detailing the study’s purpose and eligibility criteria. Promotional banners were also posted in common areas and facility entrances to encourage participation. Using a facility-based convenience sampling strategy, all eligible FHWs present during RA visits were approached directly between clinical duties. FHWs interested in participation were brought to a private area, where they were further informed about the study. Recruitment at each facility continued until no additional eligible or willing participants remained. Recruitment concluded once the target sample size of 250 FHWs was reached.

### Consent Procedures

Before enrollment, RAs provided participants with an information sheet and thoroughly explained the study objectives, time commitment, and any potential risks and benefits. FHWs were given adequate time to review the participant information sheet and the consent form, or if preferred, it was read aloud to them by the RA. These forms emphasized that participation was voluntary and that refusal or withdrawal would have no effect on employment or benefits. Participants had the opportunity to ask clarifying questions. Individuals requesting more time were invited to return at a later time. Those who agreed to participate provided written informed consent. Signed consent forms were stored securely and separately from the survey data to maintain anonymity. A total of 250 FHWs consented and completed the survey.

### Data Collection

Data were collected between January 30th, 2025, and February 7th, 2025, across 18 healthcare facilities in the Ada East district (Figure 1). Prior to launch, the survey was piloted with 10 healthcare workers at a nearby hospital to assess the clarity of questions, the flow of the survey, potential technical issues with tablets, and survey completion time. Of note, several items were adapted from previously validated global health and outbreak preparedness instruments (47–52). Pilot feedback led to minor wording revisions to improve clarity within the local context; for example, clarifying medical facility names and adding local examples of zoonotic diseases were incorporated.

Trained Ghanaian RAs, fluent in English, administered the structured, interviewer-facilitated survey in private settings to ensure comfort and confidentiality. RAs read each question aloud, clarified items as needed, and entered responses directly into REDCap using tablet computers. All RAs had prior survey administration experience and completed a one-day training on the study protocol, questionnaire content, REDCap navigation, and consent procedures. Surveys lasted 45 – 60 minutes and were conducted in English, the working language of Ghanaian health facilities. REDCap enabled offline data entry with encrypted synchronization, automatic timestamping, and completeness checks to minimize missing responses. No personal identifiers were stored with survey data.

### Survey Instrument and Measures

The survey instrument was a structured questionnaire that assessed sociodemographics, knowledge of zoonotic diseases —including topics on EVD and One Health— attitudes toward zoonoses and One Health, and zoonotic disease preventative practices. All questions were closed-ended (e.g., yes/no, multiple choice, Likert-scale), with open-text fields for any “Other” responses. Composite knowledge, attitude, and practices (KAP) scores were derived as described below.

#### Participant Characteristics

Respondents reported sociodemographic, professional, training, and information access characteristics, later evaluated as candidate predictors of KAP. Age was recorded in years (analyzed continuously and by tertiles), sex at birth was coded as a binary factor, and education was grouped as senior secondary/technical (SSS/SHS) versus higher education. Respondents indicated their primary facility type (e.g., Community-based Health Planning and Services (CHPS) compound, Health Center, or District/Regional Hospital). Occupational categories included Nurses, Midwives, other clinical staff (e.g., medical practitioners, laboratory staff, EMS personnel, pharmacists), or administrative and support staff. Job functions captured included provision of clinical services, clinical support services, and preventative health services. Years of experience in the health sector and years treating infectious diseases were categorized (≤ 5 years vs > 5 years).

Training history captured whether participants had received formal infectious disease training, and if so, the main provider of training (facility-based, government, NGO, other/none) and covered topics (e.g., COVID-19, EVD, rabies, monkeypox, etc.) were recorded. Information access was measured across electronic (e.g., mobile phones, smartphones, social media, internet), conventional (e.g., newspapers, radio, television), and public (e.g., friends, family, community meetings) sources for zoonotic-disease and climate-change information. The frequency of seeking climate-change information was rated on a five-point Likert scale. Variables with < 5% prevalence or near-zero variance were excluded from modeling. Cases with missing predictors were omitted list-wise from regression analyses.

#### Knowledge of Zoonotic Diseases

Overall knowledge was measured via 23 items derived from WHO and CDC fact sheets (3,48,53–55). Content covered the definition of zoonotic disease, identification of endemic zoonoses, transmission routes, EVD facts, and One Health principles. Correct answers were scored 1 point; incorrect or “Don’t know” answers scored 0. Total knowledge scores were the sum of points from all knowledge items, with a higher score indicating greater factual knowledge about zoonotic diseases, EVD, and One Health.

#### Attitudes Towards Zoonotic Diseases

Attitudes were assessed using 22 statements across six domains: perceived personal risk, stigma toward infected individuals, confidence in system preparedness, value of intersectoral collaboration, perceived barriers to zoonotic disease prevention, and personal preparedness for zoonotic disease management. Items used dichotomous (agree/disagree) or Likert scales, and included both positively and negatively phrased statements. We coded attitude responses such that favorable attitudes yielded a higher score. For positively phrased statements, an answer of “Agree” or “Strongly agree” was treated as a positive attitude (scored 1), while “Neutral”, “Disagree”, or “Strongly disagree” was scored 0. Reverse coding was used for negatively worded items. We then summed the points across all attitude items to obtain each participant’s total attitude score. Sub-scores were calculated for stigma-related zoonotic disease attitudes (10 items, positive attitude ≥7) and zoonotic disease preparedness and confidence (12 items, positive attitude ≥8).

#### Practices Related to Zoonotic Diseases

Zoonotic disease prevention and outbreak response practices were assessed through a 9-item frequency-based scale on personal protective equipment (PPE) use, patient education, reporting, and waste disposal. Only “Always” responses were scored as 1; a response indicating a lapse or lack of practice (e.g., Sometimes, Never) was scored as 0. Additional binary items assessed participation in zoonotic disease-specific training and cross-sector collaboration, with “Yes” scored as 1, indicative of good practices. Those answering “No” were scored as 0. Total practice scores reflected cumulative zoonotic disease prevention and control behaviors.

#### KAP Scoring and Classification

For each participant, domain-specific KAP scores were calculated by summing up item responses. These totals were converted into percentage scores based on the maximum possible score per domain. Consistent with Bloom’s taxonomy style criteria widely adopted in KAP research (56–61), percentage scores were dichotomized: participants scoring >60% were classified as having “good knowledge”, “positive attitudes”, or “good practices” within the designated KAP domain, while those scoring ≤60% were categorized as having “poor” or “negative” KAP. These binary indicators were used in descriptive analyses and regression models to identify significant predictors.

### Statistical Analysis

Data were analyzed in R Studio v4.3.1. Continuous variables were presented as means ± standard deviations; categorical variables were shown as frequencies and percentages. Composite KAP scores were calculated by summing item responses, converting totals to percentages, and dichotomizing using Bloom’s taxonomy (> 60% = “Good/Positive”; ≤ 60% = “Poor/Negative” KAP). Facility locations were geocoded and visualized using ArcGIS Online v3.2 to display participant distributions across Ada East.

To identify predictors of good knowledge and practices, univariate and multivariable logistic regression models were fitted. Candidate predictors were defined a priori and included sociodemographic characteristics (e.g., sex, age), professional factors (e.g., facility type, occupational role, years of experience, infectious disease care), training history (e.g., provider, content), and information sources (e.g., type, frequency). Univariate models produced crude odds ratios (ORs) and 95% confidence intervals (CIs), followed by multivariable models yielding adjusted odds ratios (AORs) with 95% CIs. Statistical significance was assessed using two-sided Wald chi-squared tests (α = 0.05). Model diagnostics included checks for convergence warnings and separation. Due to the small number of participants with positive attitudes (n = 17, 6.8 %), multivariable modeling was not performed to avoid overfitting.

#### Ethical considerations

This study protocol was approved by the Ghana Health Service Ethical Review Committee (GHS-ERC: 024/08/24) and the Pennsylvania State University Institutional Review Board (STUDY00025492). Additional permission was obtained from the management of each participating health facility. All participants provided written informed consent after receiving a clear explanation of the study’s risks and benefits. The study posed minimal risk beyond the time required to complete the survey, and simple language and the opportunity to ask questions ensured full understanding from participants. Participant privacy was rigorously protected. Surveys were anonymous, with no personal identifiers in the final data set. A separate encrypted file linking unique study IDs to contact information —accessible only to principal investigators— was maintained for optional focus group follow-up.

## Results

### Participant Sociodemographic, Professional, Training, and Information Characteristics

A total of 250 FHWs in Ada East were surveyed (Table 1, Fig 1). Most were women (73.2%) with a mean age of 32.8 years (SD = 6.4), and nearly all (95.6%) had completed tertiary or higher education. Participants primarily worked in district or regional hospitals (46.4%) and health centers (44.8%), with 8.8% from CHPS compounds. The majority were nurses (61.2%), followed by midwives (18.0%), other clinical staff (16.4%), and administrative support staff (4.4%). Respondents averaged 6.0 years of experience (SD = 5.6). Most provided direct clinical services (98.0%), participated in preventative services (91.6%), and 37.2% delivered clinical support services. Additionally, 63.4% reported actively diagnosing or treating infectious diseases (Table 1).

**Table 1.**
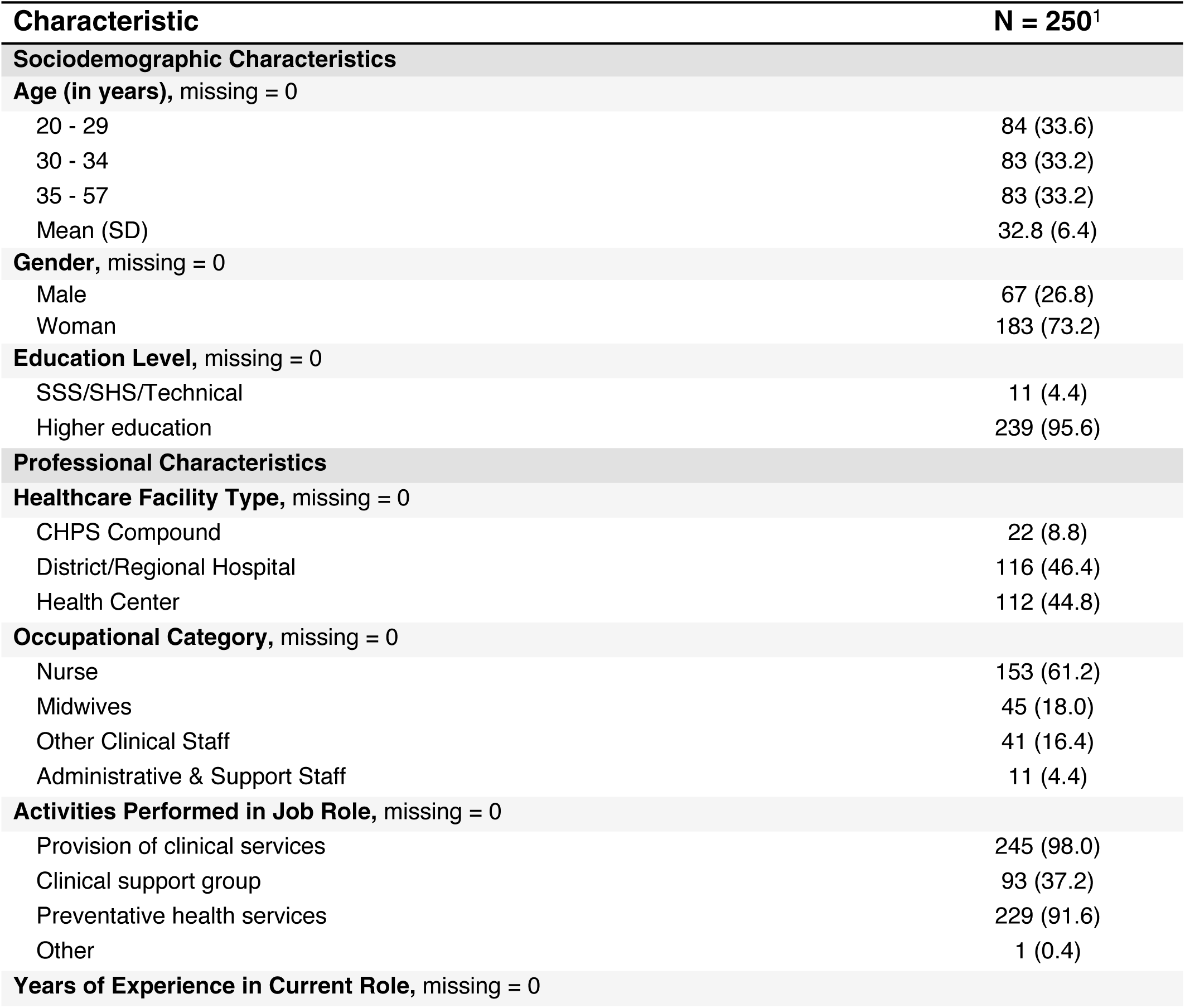
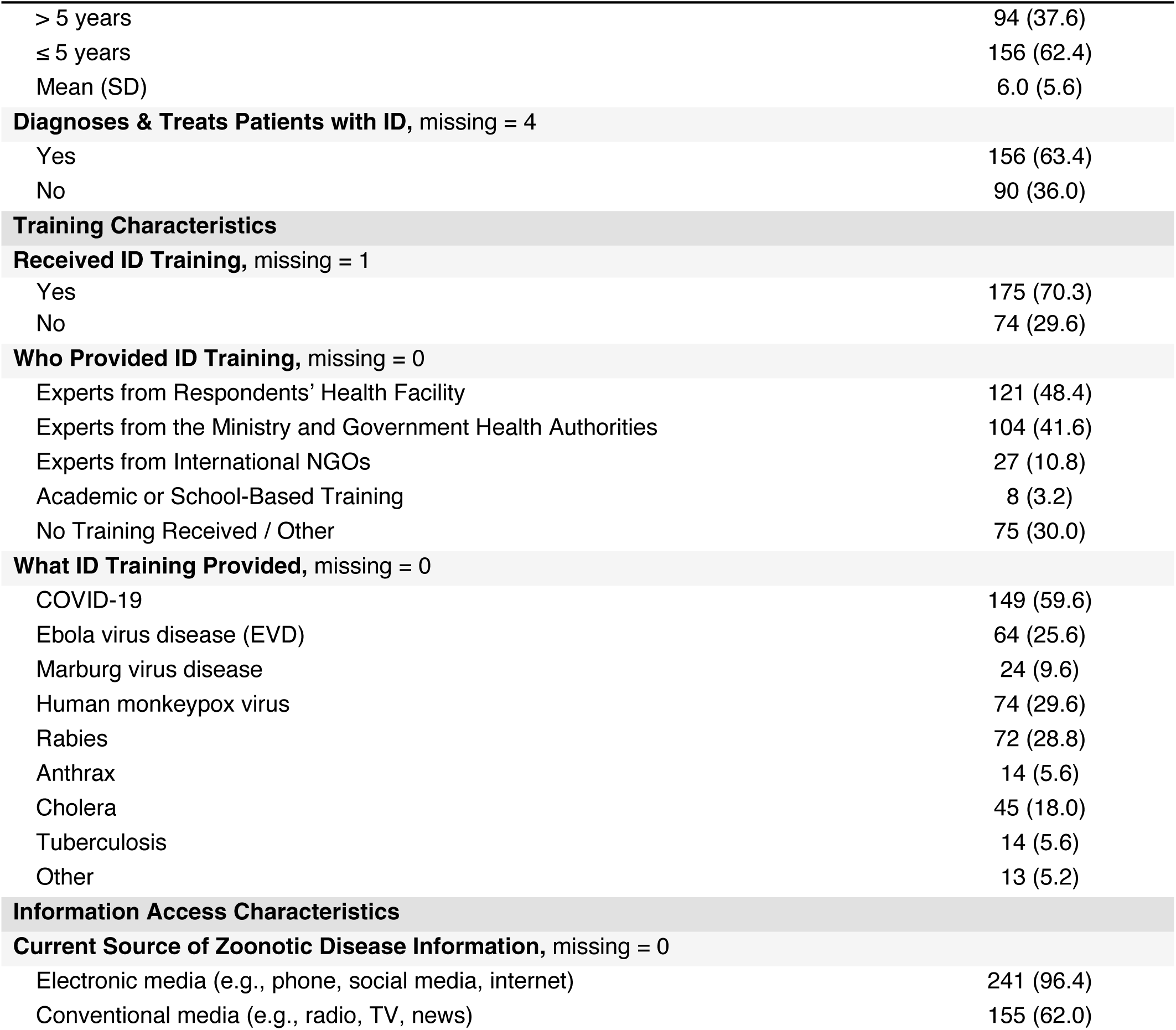
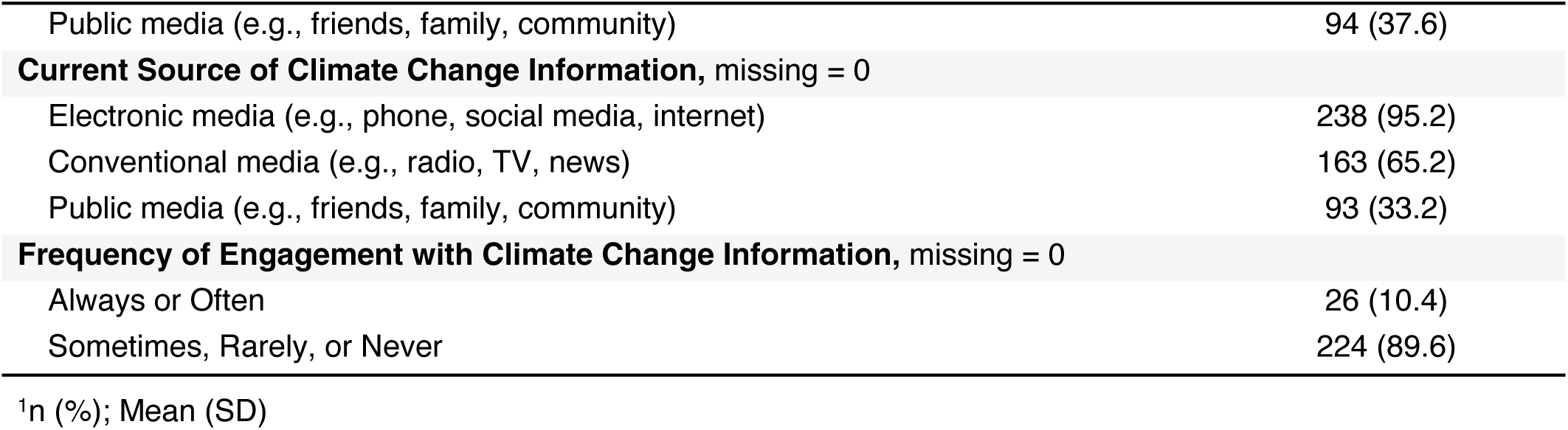
Sociodemographic, Professional, Training, and Information-Access Characteristics of Frontline Healthcare Workers (N = 250) This table presents descriptive statistics for the study sample of frontline healthcare workers (N = 250). Variables include sociodemographic characteristics (age, sex, education), professional role (facility type, occupation, years of experience), and routine job activities. Training-related variables reflect prior exposure to infectious disease (ID) training, including content and training provider. Sources of information on zoonotic diseases and climate change are also included. Categorical variables are reported as count and percent; continuous variables as mean and standard deviation. Multi-response items (e.g., training topics and information sources) are not mutually exclusive and may exceed 100%. Abbreviations: CHPS = Community-based Health Planning and Services; SSS/SHS = senior secondary/technical; ID = infectious disease; NGO = non-governmental organization; EVD = Ebola virus disease.

| Characteristic | N = 250<sup>1</sup> |
| --- | --- |
| **Sociodemographic Characteristics** |  |
| **Age (in years), missing = 0** |  |
| 20 - 29 | 84 (33.6) |
| 30 - 34 | 83 (33.2) |
| 35 - 57 | 83 (33.2) |
| Mean (SD) | 32.8 (6.4) |
| **Gender, missing = 0** |  |
| Male | 67 (26.8) |
| Woman | 183 (73.2) |
| **Education Level, missing = 0** |  |
| SSS/SHS/Technical | 11 (4.4) |
| Higher education | 239 (95.6) |
| **Professional Characteristics** |  |
| **Healthcare Facility Type, missing = 0** |  |
| CHPS Compound | 22 (8.8) |
| District/Regional Hospital | 116 (46.4) |
| Health Center | 112 (44.8) |
| **Occupational Category, missing = 0** |  |
| Nurse | 153 (61.2) |
| Midwives | 45 (18.0) |
| Other Clinical Staff | 41 (16.4) |
| Administrative & Support Staff | 11 (4.4) |
| **Activities Performed in Job Role, missing = 0** |  |
| Provision of clinical services | 245 (98.0) |
| Clinical support group | 93 (37.2) |
| Preventative health services | 229 (91.6) |
| Other | 1 (0.4) |
| **Years of Experience in Current Role, missing = 0** |  |

| **Characteristic** | **N = 250<sup>1</sup>** |
| --- | --- |
| > 5 years | 94 (37.6) |
| ≤ 5 years | 156 (62.4) |
| Mean (SD) | 6.0 (5.6) |
| **Diagnoses & Treats Patients with ID, missing = 4** |  |
| Yes | 156 (63.4) |
| No | 90 (36.0) |
| **Training Characteristics** |  |
| **Received ID Training, missing = 1** |  |
| Yes | 175 (70.3) |
| No | 74 (29.6) |
| **Who Provided ID Training, missing = 0** |  |
| Experts from Respondents' Health Facility | 121 (48.4) |
| Experts from the Ministry and Government Health Authorities | 104 (41.6) |
| Experts from International NGOs | 27 (10.8) |
| Academic or School-Based Training | 8 (3.2) |
| No Training Received / Other | 75 (30.0) |
| **What ID Training Provided, missing = 0** |  |
| COVID-19 | 149 (59.6) |
| Ebola virus disease (EVD) | 64 (25.6) |
| Marburg virus disease | 24 (9.6) |
| Human monkeypox virus | 74 (29.6) |
| Rabies | 72 (28.8) |
| Anthrax | 14 (5.6) |
| Cholera | 45 (18.0) |
| Tuberculosis | 14 (5.6) |
| Other | 13 (5.2) |
| **Information Access Characteristics** |  |
| **Current Source of Zoonotic Disease Information, missing = 0** |  |
| Electronic media (e.g., phone, social media, internet) | 241 (96.4) |
| Conventional media (e.g., radio, TV, news) | 155 (62.0) |
| Public media (e.g., friends, family, community) | 94 (37.6) |
| **Current Source of Climate Change Information, missing = 0** |  |
| Electronic media (e.g., phone, social media, internet) | 238 (95.2) |
| Conventional media (e.g., radio, TV, news) | 163 (65.2) |
| Public media (e.g., friends, family, community) | 93 (33.2) |
| **Frequency of Engagement with Climate Change Information, missing = 0** |  |
| Always or Often | 26 (10.4) |
| Sometimes, Rarely, or Never | 224 (89.6) |

<sup>1</sup>n (%); Mean (SD)

Regarding training, 70.3% had prior infectious disease training, primarily delivered by in-facility experts (48.4%) or government health authorities (41.6%). Common training topics included COVID-19 (59.6%), human monkeypox (29.6%), rabies (28.8%), EVD (25.6%), and cholera (18.0%), among others less frequently covered (Table 1).

Nearly all FHWs used electronic sources for zoonotic (96.4%) and climate-related (95.2%) health information (Table 1). Conventional media use was also common (62.0% for zoonoses; 65.2% for climate change), meanwhile public networks were less frequently cited (37.6% for zoonoses; 33.2% for climate change). Climate change information engagement was generally low, with 89.6% reporting infrequent access (i.e., sometimes, rarely, or never).

### Frontline Healthcare Workers’ Knowledge of Zoonotic Diseases, EVD, and One Health

Table 2 summarizes knowledge scores across three domains: zoonotic disease identification, Ebola-specific knowledge, and One Health. Overall, 79.2% of participants scored >60% on the total knowledge scale, meeting the threshold for “good knowledge” on zoonoses identification, EVD, and One Health; 20.8% scored below this cutoff. In this study, 77.6% scored below the 60% threshold, indicating limited recognition of common zoonoses. While EVD (98.0%) and rabies (91.6%) were widely recognized as zoonotic diseases, nearly half identified influenza (49.6%), with fewer recognizing salmonellosis (23.6%) or Lyme disease (18.0%).

**Table 2.**
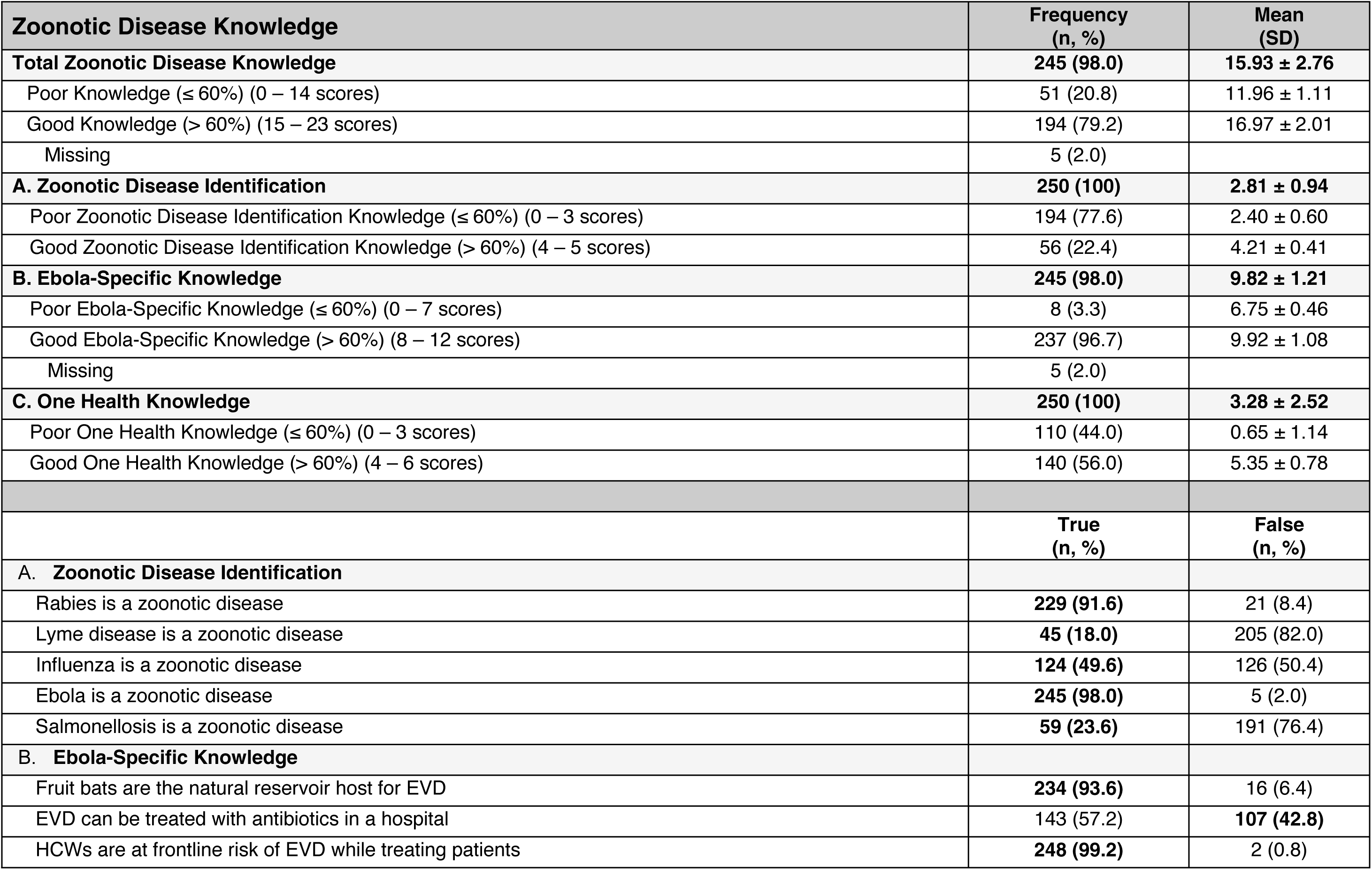
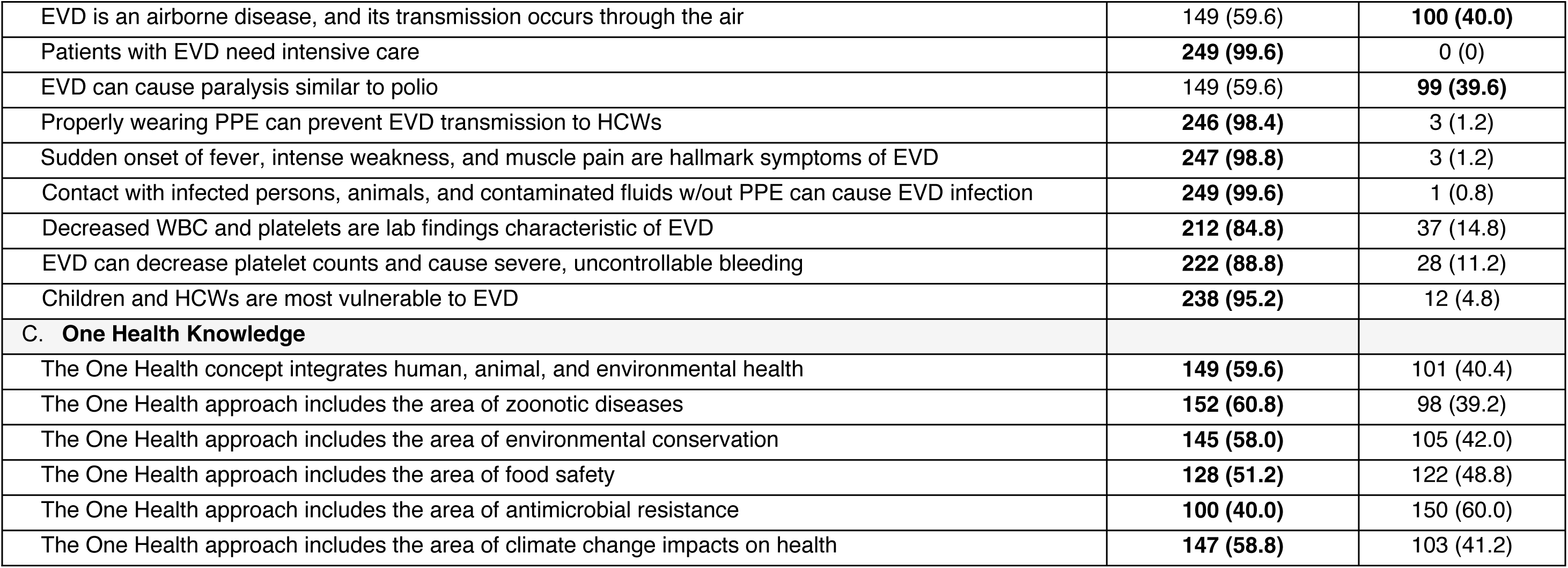
Knowledge of Zoonotic Diseases, EVD, and One Health Among Frontline Healthcare Workers (N = 250) Bolded values in the item section indicate the correct response. Composite knowledge scores were calculated across three domains: zoonotic disease identification (5 items), Ebola virus disease-specific knowledge (12 items), and One Health (6 items), for a maximum total score of 23. Each correct response was scored as 1. Total knowledge scores >60% threshold (>14/23) were classified as “good knowledge,” and ≤60% as “poor knowledge.” Five participants (2.0%) had one or more missing responses, all within the Ebola knowledge domain. Abbreviations: EVD = Ebola virus disease, PPE = personal protective equipment, HCW = health care worker.

| **Zoonotic Disease Knowledge** | **Frequency**<br>**(n, %)** | **Mean**<br>**(SD)** |
| --- | --- | --- |
| **Total Zoonotic Disease Knowledge** | **245 (98.0)** | **15.93 ± 2.76** |
| Poor Knowledge ($\leq 60\%$) (0 – 14 scores) | 51 (20.8) | 11.96 ± 1.11 |
| Good Knowledge ($> 60\%$) (15 – 23 scores) | 194 (79.2) | 16.97 ± 2.01 |
| Missing | 5 (2.0) |  |
| **A. Zoonotic Disease Identification** | **250 (100)** | **2.81 ± 0.94** |
| Poor Zoonotic Disease Identification Knowledge ($\leq 60\%$) (0 – 3 scores) | 194 (77.6) | 2.40 ± 0.60 |
| Good Zoonotic Disease Identification Knowledge ($> 60\%$) (4 – 5 scores) | 56 (22.4) | 4.21 ± 0.41 |
| **B. Ebola-Specific Knowledge** | **245 (98.0)** | **9.82 ± 1.21** |
| Poor Ebola-Specific Knowledge ($\leq 60\%$) (0 – 7 scores) | 8 (3.3) | 6.75 ± 0.46 |
| Good Ebola-Specific Knowledge ($> 60\%$) (8 – 12 scores) | 237 (96.7) | 9.92 ± 1.08 |
| Missing | 5 (2.0) |  |
| **C. One Health Knowledge** | **250 (100)** | **3.28 ± 2.52** |
| Poor One Health Knowledge ($\leq 60\%$) (0 – 3 scores) | 110 (44.0) | 0.65 ± 1.14 |
| Good One Health Knowledge ($> 60\%$) (4 – 6 scores) | 140 (56.0) | 5.35 ± 0.78 |
|  | **True**<br>**(n, %)** | **False**<br>**(n, %)** |
| **A. Zoonotic Disease Identification** |  |  |
| Rabies is a zoonotic disease | **229 (91.6)** | 21 (8.4) |
| Lyme disease is a zoonotic disease | **45 (18.0)** | 205 (82.0) |
| Influenza is a zoonotic disease | **124 (49.6)** | 126 (50.4) |
| Ebola is a zoonotic disease | **245 (98.0)** | 5 (2.0) |
| Salmonellosis is a zoonotic disease | **59 (23.6)** | 191 (76.4) |
| **B. Ebola-Specific Knowledge** |  |  |
| Fruit bats are the natural reservoir host for EVD | **234 (93.6)** | 16 (6.4) |
| EVD can be treated with antibiotics in a hospital | 143 (57.2) | **107 (42.8)** |
| HCWs are at frontline risk of EVD while treating patients | **248 (99.2)** | 2 (0.8) |
| EVD is an airborne disease, and its transmission occurs through the air | 149 (59.6) | **100 (40.0)** |
| Patients with EVD need intensive care | **249 (99.6)** | 0 (0) |
| EVD can cause paralysis similar to polio | 149 (59.6) | **99 (39.6)** |
| Properly wearing PPE can prevent EVD transmission to HCWs | **246 (98.4)** | 3 (1.2) |
| Sudden onset of fever, intense weakness, and muscle pain are hallmark symptoms of EVD | **247 (98.8)** | 3 (1.2) |
| Contact with infected persons, animals, and contaminated fluids w/out PPE can cause EVD infection | **249 (99.6)** | 1 (0.8) |
| Decreased WBC and platelets are lab findings characteristic of EVD | **212 (84.8)** | 37 (14.8) |
| EVD can decrease platelet counts and cause severe, uncontrollable bleeding | **222 (88.8)** | 28 (11.2) |
| Children and HCWs are most vulnerable to EVD | **238 (95.2)** | 12 (4.8) |
| **C. One Health Knowledge** |  |  |
| The One Health concept integrates human, animal, and environmental health | **149 (59.6)** | 101 (40.4) |
| The One Health approach includes the area of zoonotic diseases | **152 (60.8)** | 98 (39.2) |
| The One Health approach includes the area of environmental conservation | **145 (58.0)** | 105 (42.0) |
| The One Health approach includes the area of food safety | **128 (51.2)** | 122 (48.8) |
| The One Health approach includes the area of antimicrobial resistance | **100 (40.0)** | 150 (60.0) |
| The One Health approach includes the area of climate change impacts on health | **147 (58.8)** | 103 (41.2) |

Among the 245 respondents with complete Ebola-specific knowledge data, 96.7% were classified as having good Ebola-specific knowledge (scored >60% in this subdomain). Nearly all participants recognized high occupational risk for healthcare workers handling zoonotic diseases (98.0%), the importance of PPE in preventing transmission (98.4%), hallmark symptoms of EVD (98.8%), and EVD transmission routes (99.6%). However, misconceptions were common— 57.2% incorrectly believed antibiotics could treat EVD, and 59.6% thought EVD was an airborne disease or could cause paralysis like polio.

For the One Health knowledge subdomain consisting of six items, 56.0% of respondents scored >60%, classified as having “good knowledge” of One Health principles. Most respondents correctly defined One Health as integrating human, animal, and environmental health (59.6%). Awareness of the broader components of One Health varied, with the majority of participants correctly identifying the principle’s inclusion of zoonotic diseases (60.8%), climate change impacts on health (58.8%), environmental conservation (58.0%), and food safety (51.2%), though fewer recognized antimicrobial resistance (40.0%) as part of the approach (Table 2).

### Frontline Healthcare Workers’ Attitudes Toward Zoonotic Diseases and Outbreak Preparedness

FHW’s overall attitudes —reflecting perceptions of personal and institutional preparedness, confidence in managing zoonotic diseases, and stigma— were predominantly negative (Table 3). Only 6.8% of participants scored >60% (≥14 points), meeting the threshold for “positive attitudes.” The remaining 93.2% of participants fell below this threshold, indicating widespread concerns about personal safety, institutional readiness, and community-level stigma.

**Table 3.** Attitudes Toward Zoonotic Diseases and Preparedness Among Frontline Healthcare Workers (N = 250) Bolded values in the item sections reflect positive attitudes. Responses were coded as binary, with favorable (i.e., positive) responses assigned a score of 1, and all others scored as 0. The full-scale attitude score (22 items) was used to derive “Positive Attitudes” ( >60% threshold; >14 points out of 22). Attitude subscales included: (A) Stigma-related Zoonotic Disease Attitudes (10 items; positive ≥7), and (B) Preparedness and Confidence Attitudes (12 items; high ≥8). Stigma-related attitudes were assessed through agreement or disagreement with statements reflecting stigma-related beliefs toward infected individuals. Preparedness and confidence were assessed through perceived risk, institutional readiness, personal confidence, and recognition of structural barriers to zoonotic disease prevention. No missing responses were recorded. Abbreviations: EVD = Ebola virus disease.

| **Zoonotic Disease Attitudes** | **Frequency**<br>**(n, %)** | **Mean**<br>**(SD)** |
| --- | --- | --- |
| **Overall Zoonotic Disease Attitudes** | **250 (100)** | **9.30 ± 2.54** |
| Negative Attitudes ($\leq 60\%$) (0 – 13 scores) | 233 (93.2) | 8.91 ± 2.17 |
| Positive Attitudes ($> 60\%$) (14 – 22 scores) | 17 (6.8) | 14.59 ± 0.71 |
| **A. Stigma-Related Zoonotic Disease Attitudes** | **250 (100)** | **4.11 ± 1.74** |
| Negative Attitudes ($\leq 60\%$) (0 – 6 scores) | 233 (93.2) | 3.88 ± 1.57 |
| Positive Attitudes ($> 60\%$) (7 – 10 scores) | 17 (6.8) | 7.23 ± 0.56 |
| **B. Zoonotic Disease Preparedness & Confidence Attitudes** | **250 (100)** | **5.19 ± 1.58** |
| Low preparedness & confidence ($\leq 60\%$) (0 – 7 scores) | 208 (83.2) | 4.67 ± 1.09 |
| High preparedness & confidence ($> 60\%$) (8 – 12 scores) | 42 (16.8) | 7.79 ± 0.92 |
|  | **Agree**<br>**(n, %)** | **Disagree**<br>**(n, %)** |
| **A. Stigma-Related Zoonotic Disease Attitudes** |  |  |
| Individuals infected with a zoonotic disease should be legally separated from others to protect the public | 241 (96.4) | **9 (3.6)** |
| Names of individuals infected with a zoonotic disease should be made public so others can avoid them | 33 (13.2) | **217 (86.8)** |
| Patients with a zoonotic disease (e.g., EVD) should be treated separately from other patients | 241 (96.4) | **9 (3.6)** |
| I would feel uncomfortable if my patients, family, or friends were infected with a zoonotic disease | 204 (81.6) | **46 (18.4)** |
| Families infected with a zoonotic disease (e.g., EVD) should be isolated from the community | 185 (74.0) | **65 (26.0)** |
| I feel uncomfortable working in a health center that treats patients with zoonotic disease (e.g., EVD) | 88 (35.2) | **162 (64.8)** |
| Countries affected by zoonotic diseases should close their airports and seaports to prevent transmission | 212 (84.8) | **38 (15.2)** |
| Zoonotic diseases, like EVD, are only a threat to Africa, not to the rest of the world | 68 (27.2) | **182 (72.8)** |
| HIV/AIDS is a bigger threat than zoonotic diseases (e.g., EVD) in West Africa (in the study region, Ghana) | 76 (30.4) | **174 (69.6)** |
| I am more concerned about my own life than treating patients with zoonotic diseases, like EVD | 125 (50.0) | **125 (50.0)** |
| **B. Zoonotic Disease Preparedness & Confidence Attitudes** |  |  |
| I am concerned about the risk of a zoonotic disease outbreak in my community, in Ghana, and in the region | 244 (97.6) | **6 (2.4)** |
| My health center is adequately prepared to handle any zoonotic disease outbreak | **72 (28.8)** | 178 (71.2) |
| Ghana and the region are adequately prepared to handle any zoonotic disease outbreak | **126 (50.4)** | 124 (49.6) |
| I am confident in my ability to identify symptoms of zoonotic diseases in patients | **237 (94.8)** | 13 (5.2) |
| I am confident in my health center's capacity to manage a zoonotic disease outbreak | **195 (78.0)** | 55 (22.0) |
| Collaboration between human health, animal health, and environmental health sectors is beneficial | **244 (97.6)** | 6 (2.4) |
| The One Health approach is effective in preventing zoonotic disease outbreaks | **228 (91.2)** | 22 (8.8) |
| I perceive the need for more frequent training sessions as a barrier to zoonotic disease prevention | **20 (8.0)** | 230 (92.0) |
| I perceive the lack of access to updated guidelines as a barrier to zoonotic disease prevention | **34 (13.6)** | 216 (86.4) |
| I perceive limited interdisciplinary workshops as a barrier to zoonotic disease prevention | **47 (18.8)** | 203 (81.2) |
| I perceive challenges in collaborating with other sectors as a barrier to zoonotic disease prevention | **50 (20.0)** | 200 (80.0) |
| I perceive insufficient information on One Health as a barrier to zoonotic disease prevention | **39 (15.6)** | 211 (84.4) |

Stigma-related attitudes were common. Agreement with restrictive measures was particularly high, where 96.4% supported legal separation of infected patients from the public, 74.0% favored isolating infected families, and 84.8% endorsed border closures. However, 86.8% opposed publicly disclosing the names of infected individuals to the public (Table 3). Half of the participants prioritized personal safety over treating infected patients, and 35.2% were uncomfortable working in facilities managing EVD. Moreover, 30.4% perceived HIV/AIDS as a greater health concern than zoonotic diseases in West Africa, and 27.2% viewed EVD as a threat exclusive to only Africa.

Attitudes of low preparedness and confidence were also prevalent (Table 3). While 97.6% were concerned about local outbreaks and 94.8% felt confident in identifying symptoms of zoonoses, fewer trusted institutional readiness. That is, only 28.8% believed their facility was prepared, and 50.4% believed Ghana and the West African region were equipped for adequate zoonotic disease outbreak response.

Despite these concerns, support for the One Health approach was strong. Particularly, 97.6% agreed that cross-sector collaboration is beneficial, and 91.2% believed it helps to prevent outbreaks. Nonetheless, structural barriers to One Health operationalization were frequently reported, including cross-sector collaboration challenges (20.0%), limited One Health information (15.6%), absence of interdisciplinary workshops (18.8%), and poor access to updated zoonotic disease management guidelines (13.6%).

### Frontline Healthcare Workers’ Zoonotic Disease Prevention and Control Practices

Total practice scores —capturing both individual-level and institutional-level zoonotic disease prevention and control practices— ranged from 0 to 9, with a mean score of 5.35 (SD = 1.54). Based on the >60% threshold (>6 correct items), 47.2% of participants demonstrated “good practices,” while 52.8% fell below this (Table 4).

**Table 4.** Zoonotic Disease Prevention and Control Practices of Frontline Healthcare Workers (N = 250) Bolded values indicate responses reflecting good zoonotic disease prevention and control practices at the individual and institutional levels. Nine practice items were recoded into binary indicators, with “Yes” representing a “good practice” (scored 1) and “No” representing a “poor practice”. Participants scoring >6 out of 9 (>60%) were classified as having “Good Practices,” while those scoring ≤5 were classified as having “Poor Practices.” Items assessed individual-level behaviors (e.g., PPE use, disease reporting, patient education, vaccination guidance, biomedical waste management), institutional engagement (e.g., institutional protocols and training), and multisectoral collaboration related to zoonotic disease prevention and response.

| **Zoonotic Disease Practices** | **Frequency**<br>**(n, %)** | **Mean**<br>**(SD)** |
| --- | --- | --- |
| **Overall Zoonotic Disease Practices** | **250 (100)** | **5.35 ± 1.54** |
| Poor Practices (≤ 60%) (0 – 5 scores) | 132 (52.8) | 4.20 ± 0.98 |
| Good Practices (> 60%) (6 – 9 scores) | 118 (47.2) | 6.64 ± 0.87 |
|  | **Yes**<br>**(n, %)** | **No**<br>**(n, %)** |
| **Zoonotic Disease Practices** |  |  |
| Always use PPE when caring for patients suspected or confirmed to have a zoonotic disease | **170 (68.0)** | 80 (32.0) |
| Always report any suspected zoonotic disease cases, like Ebola, to the appropriate health authorities | **232 (92.8)** | 18 (7.2) |
| Always educate patients about the signs, symptoms, and prevention methods of zoonotic diseases | **183 (73.2)** | 67 (26.8) |
| Always advise patients to take vaccines to help prevent zoonotic diseases | **205 (82.0)** | 45 (18.0) |
| Always ensure that all waste generated while treating patients with zoonotic disease is disposed of according to safety guidelines | **233 (93.2)** | 17 (6.8) |
| Received formal training on zoonotic disease prevention and control in the past year | **65 (26.0)** | 185 (74.0) |
| There is a protocol in place for handling zoonotic disease outbreaks in my health facility | **180 (72.0)** | 70 (28.0) |
| Regularly collaborate with professionals from other sectors (e.g., veterinary, environmental) on health-related issues | **47 (18.8)** | 203 (81.2) |
| Received formal training on the One Health concept in the past year | **23 (9.2)** | 227 (90.8) |

Most participants reported engaging in individual-level prevention behaviors (Table 4). PPE use was consistent among 68.0% of respondents when caring for suspected zoonotic disease cases. Nearly all FHWs reported always notifying authorities of suspected cases (92.8%) and ensuring proper medical waste disposal (93.2%). Additionally, 73.2% regularly educated patients on zoonotic symptoms and prevention, and 82.0% advised vaccination when applicable.

Institutional training and engagement in collaborative practices were minimal (Table 4).

Only 26.0% had received formal zoonotic disease training in the past year, and just 9.2% had received One Health training. Although 72.0% reported that their health facility had zoonotic disease management and outbreak protocols, only 18.8% routinely collaborated with professionals from other sectors.

### Predictors of Zoonotic Disease Knowledge and Practices

Table 5 shows predictors of scoring >60% on the total zoonotic disease knowledge scale (classified as “good knowledge”). Compared to FHWs aged 20-29, those aged 30-34 (AOR = 0.28, 95% CI: 0.09-0.81, p=0.022) and 35-57 (AOR = 0.24, 95% CI: 0.06-0.87, p=0.032) had significantly lower odds of good knowledge on zoonoses identification, EVD, and One Health. Government-provided infectious disease training was also associated with lower odds of good knowledge (AOR = 0.37, 95% CI: 0.15–0.87, p = 0.024), as was training focused on EVD (AOR = 0.17, 95% CI: 0.05–0.57, p = 0.005). Rabies-related training significantly increased the odds of good knowledge (AOR = 3.25, 95% CI: 1.04–11.1, p = 0.050).

**Table 5.**
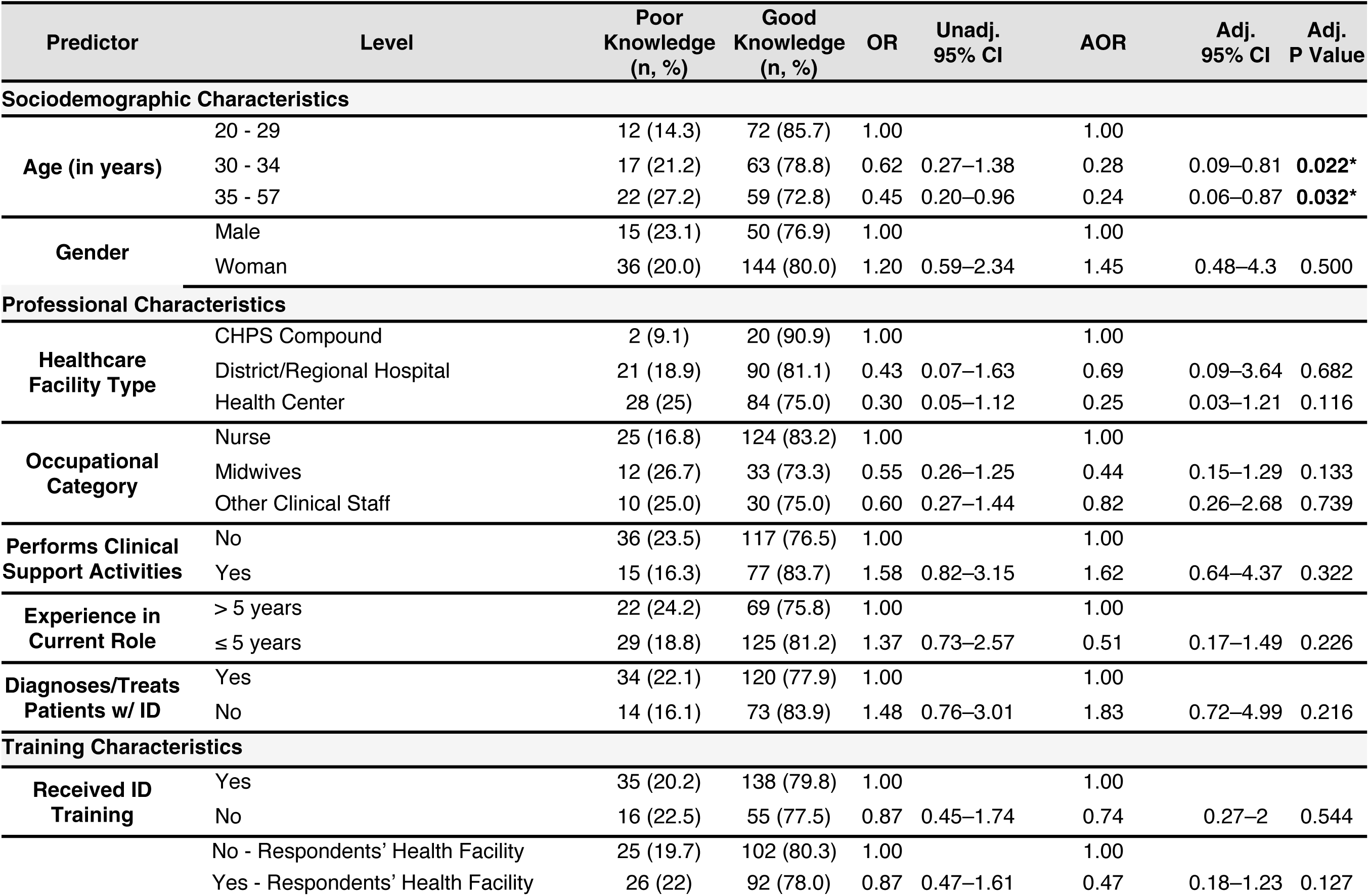
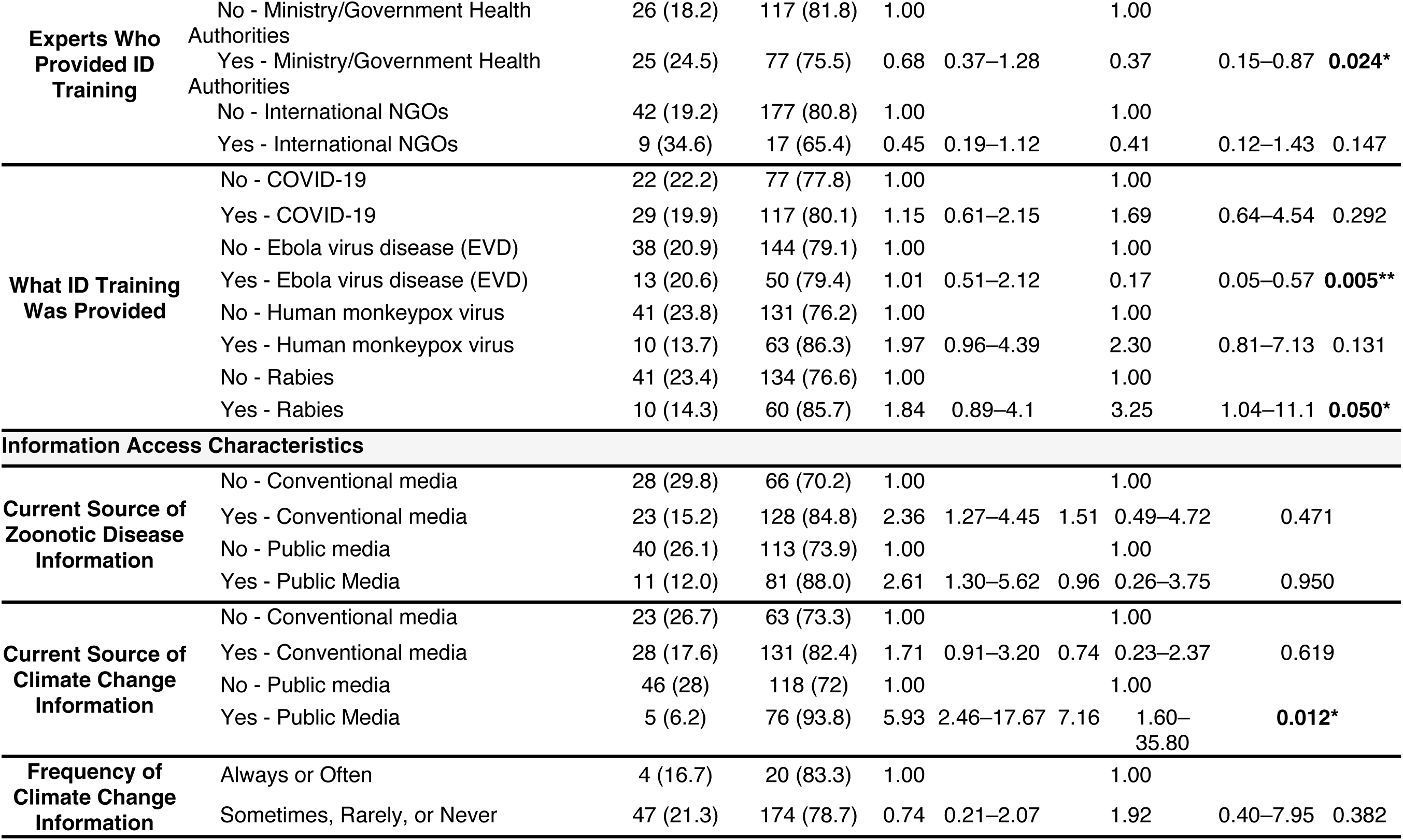
Predictors of Good Knowledge (>60%) of Zoonotic Diseases, EVD, and One Health among Frontline Healthcare Workers (N = 250) This table presents logistic regression results examining associations between sociodemographic, professional, training, and information access characteristics and “good knowledge” across the knowledge domains of zoonoses identification, EVD, and One Health. Both unadjusted odds ratios (OR) and adjusted odds ratios (AOR) are reported, each with corresponding 95% confidence intervals (CI). Values are rounded to two decimal places for ORs and CIs, and three decimal places for p-values. A significance threshold of α = 0.05 was applied; statistically significant results are bolded and indicated as follows: p < 0.05 (*), p < 0.01 (**), and p < 0.001 (***). Abbreviations: ID = infectious disease; COVID-19 = coronavirus disease 2019; EVD = Ebola virus disease; CHPS = Community-based Health Planning and Services; NGO = nongovernmental organization; Unadj. = unadjusted; Adj. = adjusted.

Table 6 summarizes predictors of scoring >60% on the zoonotic disease prevention and control practices scale (classified as “good practices”). FHWs who did not receive any infectious disease training had significantly lower odds of good practices (AOR = 0.43, 95% CI: 0.19–0.94, p = 0.036), while facility-based training was associated with higher odds of good practices (AOR = 2.77, 95% CI: 1.34–5.92, p = 0.007). Rabies-related training also significantly predicted higher odds of good practices (AOR = 2.46, 95% CI: 1.07–5.87, p = 0.037), compared to other disease-specific training on COVID-19, EVD, or Human monkeypox. FHWs who engaged with climate change information only sometimes, rarely, or never had significantly lower odds of good practices (AOR = 0.29, 95% CI: 0.09–0.86, p = 0.031).

**Table 6.**
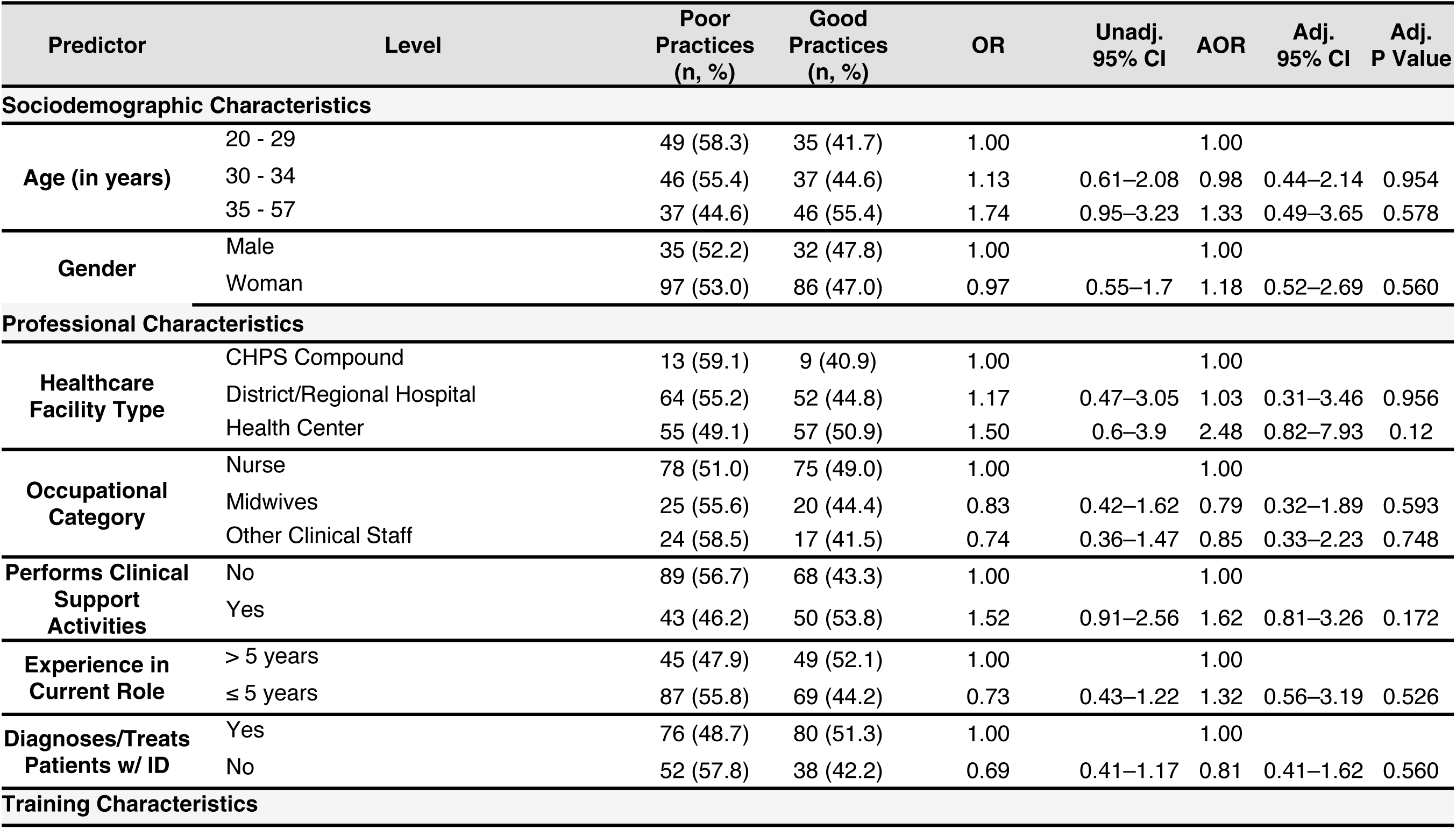
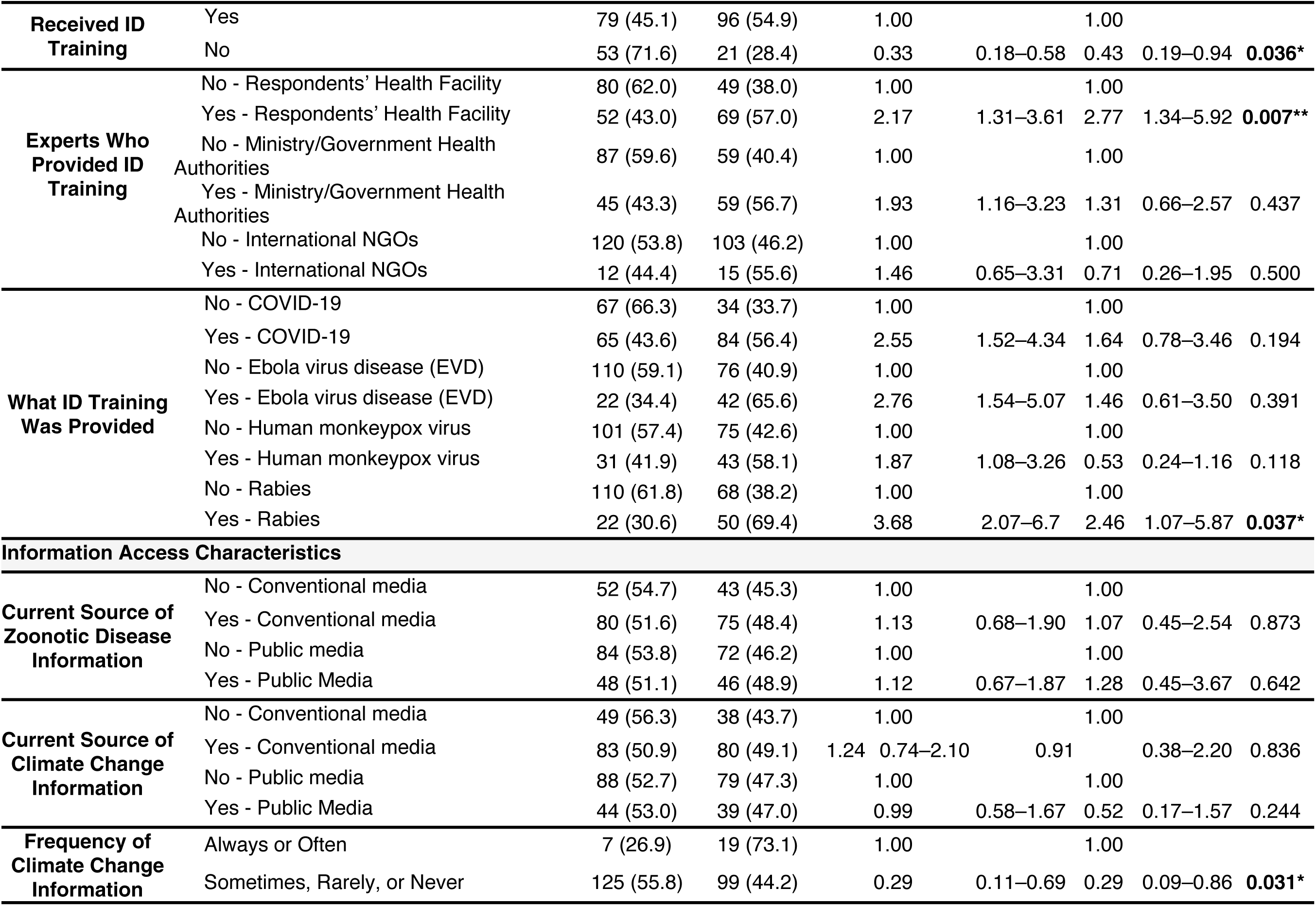
Predictors of Good Zoonotic Disease Prevention and Control Practices (>60%) among Frontline Healthcare Workers (N = 250) This table presents logistic regression results examining associations between sociodemographic, professional, training, and information access characteristics and “good practices” (>60% score) regarding zoonotic diseases prevention and control at both the individual- and institutional-level. Both unadjusted odds ratios (OR) and adjusted odds ratios (AOR) are reported, each with corresponding 95% confidence intervals (CI). Values are rounded to two decimal places for ORs and CIs, and three decimal places for p-values. A significance threshold of α = 0.05 was applied; statistically significant results are bolded and indicated as follows: p < 0.05 (*), p < 0.01 (**), and p < 0.001 (***). Abbreviations: ID = infectious disease; COVID-19 = coronavirus disease 2019; EVD = Ebola virus disease; CHPS = Community-based Health Planning and Services; NGO = nongovernmental organization; Unadj. = unadjusted; Adj. = adjusted.

| Predictor | Level | Poor Practices<br>(n, %) | Good Practices<br>(n, %) | OR | Unadj.<br>95% CI | AOR | Adj.<br>95% CI | Adj.<br>P Value |
| --- | --- | --- | --- | --- | --- | --- | --- | --- |
| **Sociodemographic Characteristics** |  |  |  |  |  |  |  |  |
| **Age (in years)** | 20 - 29 | 49 (58.3) | 35 (41.7) | 1.00 |  | 1.00 |  |  |
|  | 30 - 34 | 46 (55.4) | 37 (44.6) | 1.13 | 0.61–2.08 | 0.98 | 0.44–2.14 | 0.954 |
|  | 35 - 57 | 37 (44.6) | 46 (55.4) | 1.74 | 0.95–3.23 | 1.33 | 0.49–3.65 | 0.578 |
| **Gender** | Male | 35 (52.2) | 32 (47.8) | 1.00 |  | 1.00 |  |  |
|  | Woman | 97 (53.0) | 86 (47.0) | 0.97 | 0.55–1.7 | 1.18 | 0.52–2.69 | 0.560 |
| **Professional Characteristics** |  |  |  |  |  |  |  |  |
| **Healthcare Facility Type** | CHPS Compound | 13 (59.1) | 9 (40.9) | 1.00 |  | 1.00 |  |  |
|  | District/Regional Hospital | 64 (55.2) | 52 (44.8) | 1.17 | 0.47–3.05 | 1.03 | 0.31–3.46 | 0.956 |
|  | Health Center | 55 (49.1) | 57 (50.9) | 1.50 | 0.6–3.9 | 2.48 | 0.82–7.93 | 0.12 |
| **Occupational Category** | Nurse | 78 (51.0) | 75 (49.0) | 1.00 |  | 1.00 |  |  |
|  | Midwives | 25 (55.6) | 20 (44.4) | 0.83 | 0.42–1.62 | 0.79 | 0.32–1.89 | 0.593 |
|  | Other Clinical Staff | 24 (58.5) | 17 (41.5) | 0.74 | 0.36–1.47 | 0.85 | 0.33–2.23 | 0.748 |
| **Performs Clinical Support Activities** | No | 89 (56.7) | 68 (43.3) | 1.00 |  | 1.00 |  |  |
|  | Yes | 43 (46.2) | 50 (53.8) | 1.52 | 0.91–2.56 | 1.62 | 0.81–3.26 | 0.172 |
| **Experience in Current Role** | > 5 years | 45 (47.9) | 49 (52.1) | 1.00 |  | 1.00 |  |  |
|  | ≤ 5 years | 87 (55.8) | 69 (44.2) | 0.73 | 0.43–1.22 | 1.32 | 0.56–3.19 | 0.526 |
| **Diagnoses/Treats Patients w/ ID** | Yes | 76 (48.7) | 80 (51.3) | 1.00 |  | 1.00 |  |  |
|  | No | 52 (57.8) | 38 (42.2) | 0.69 | 0.41–1.17 | 0.81 | 0.41–1.62 | 0.560 |
| **Training Characteristics** |  |  |  |  |  |  |  |  |
| **Received ID Training** | Yes | 79 (45.1) | 96 (54.9) | 1.00 |  | 1.00 |  |  |
|  | No | 53 (71.6) | 21 (28.4) | 0.33 | 0.18–0.58 | 0.43 | 0.19–0.94 | **0.036\*** |
| **Experts Who Provided ID Training** | No - Respondents' Health Facility | 80 (62.0) | 49 (38.0) | 1.00 |  | 1.00 |  |  |
|  | Yes - Respondents' Health Facility | 52 (43.0) | 69 (57.0) | 2.17 | 1.31–3.61 | 2.77 | 1.34–5.92 | **0.007\*\*** |
|  | No - Ministry/Government Health Authorities | 87 (59.6) | 59 (40.4) | 1.00 |  | 1.00 |  |  |
|  | Yes - Ministry/Government Health Authorities | 45 (43.3) | 59 (56.7) | 1.93 | 1.16–3.23 | 1.31 | 0.66–2.57 | 0.437 |
|  | No - International NGOs | 120 (53.8) | 103 (46.2) | 1.00 |  | 1.00 |  |  |
|  | Yes - International NGOs | 12 (44.4) | 15 (55.6) | 1.46 | 0.65–3.31 | 0.71 | 0.26–1.95 | 0.500 |
| **What ID Training Was Provided** | No - COVID-19 | 67 (66.3) | 34 (33.7) | 1.00 |  | 1.00 |  |  |
|  | Yes - COVID-19 | 65 (43.6) | 84 (56.4) | 2.55 | 1.52–4.34 | 1.64 | 0.78–3.46 | 0.194 |
|  | No - Ebola virus disease (EVD) | 110 (59.1) | 76 (40.9) | 1.00 |  | 1.00 |  |  |
|  | Yes - Ebola virus disease (EVD) | 22 (34.4) | 42 (65.6) | 2.76 | 1.54–5.07 | 1.46 | 0.61–3.50 | 0.391 |
|  | No - Human monkeypox virus | 101 (57.4) | 75 (42.6) | 1.00 |  | 1.00 |  |  |
|  | Yes - Human monkeypox virus | 31 (41.9) | 43 (58.1) | 1.87 | 1.08–3.26 | 0.53 | 0.24–1.16 | 0.118 |
|  | No - Rabies | 110 (61.8) | 68 (38.2) | 1.00 |  | 1.00 |  |  |
|  | Yes - Rabies | 22 (30.6) | 50 (69.4) | 3.68 | 2.07–6.7 | 2.46 | 1.07–5.87 | **0.037\*** |
| **Information Access Characteristics** |  |  |  |  |  |  |  |  |
| **Current Source of Zoonotic Disease Information** | No - Conventional media | 52 (54.7) | 43 (45.3) | 1.00 |  | 1.00 |  |  |
|  | Yes - Conventional media | 80 (51.6) | 75 (48.4) | 1.13 | 0.68–1.90 | 1.07 | 0.45–2.54 | 0.873 |
|  | No - Public media | 84 (53.8) | 72 (46.2) | 1.00 |  | 1.00 |  |  |
|  | Yes - Public Media | 48 (51.1) | 46 (48.9) | 1.12 | 0.67–1.87 | 1.28 | 0.45–3.67 | 0.642 |
| **Current Source of Climate Change Information** | No - Conventional media | 49 (56.3) | 38 (43.7) | 1.00 |  | 1.00 |  |  |
|  | Yes - Conventional media | 83 (50.9) | 80 (49.1) | 1.24 | 0.74–2.10 | 0.91 | 0.38–2.20 | 0.836 |
|  | No - Public media | 88 (52.7) | 79 (47.3) | 1.00 |  | 1.00 |  |  |
|  | Yes - Public Media | 44 (53.0) | 39 (47.0) | 0.99 | 0.58–1.67 | 0.52 | 0.17–1.57 | 0.244 |
| **Frequency of Climate Change Information** | Always or Often | 7 (26.9) | 19 (73.1) | 1.00 |  | 1.00 |  |  |
|  | Sometimes, Rarely, or Never | 125 (55.8) | 99 (44.2) | 0.29 | 0.11–0.69 | 0.29 | 0.09–0.86 | **0.031\*** |

## Discussion

This study characterized FHWs’ KAP related to zoonotic disease, EVD, and One Health in Ada East, a peri-urban and rural district within Ghana’s Greater Accra Region. To the best of our knowledge, this is the first study in southern Ghana to assess FHW KAP on zoonoses and One Health, while also identifying professional, training, and information access factors associated with good knowledge and practices. While previous research has documented knowledge gaps in Ghana (40,41), this study adds to existing literature by examining FHWs’ attitudes and practices as well. We also examined a range of factors associated with good knowledge and practices across multiple health facilities in Ada East.

The results of this study show high awareness of high-profile diseases like EVD —nearly all respondents (98.0%) recognized Ebola as zoonotic— while recognition of more endemic zoonoses (e.g., influenza, salmonellosis) was markedly lower. This reflects findings from northern Ghana, where 75.3% identified EVD as zoonotic but only 12% recognized brucellosis, among other endemic zoonoses (41). Despite most participants demonstrating “good knowledge” of zoonoses, EVD, and One Health (79.2%), widespread fear, stigma, and minimal confidence in outbreak preparedness persisted — a disconnect consistent with past West African findings (62–65). Personal zoonotic disease prevention behaviors were more common than institutional practices, and intersectoral collaboration was limited. Few FHWs had received recent zoonotic disease training or engaged with veterinary or environmental health professionals. Compared to other regions in SSA, where intersectoral engagement is more common (66–69), our findings highlight critical gaps in Ada East’s frontline preparedness infrastructure.

### Considerations for Ghana’s One Health and Zoonotic Disease Preparedness Landscape

The KAP gaps identified in this study must be understood within Ghana’s evolving One Health strategy and outbreak preparedness landscape. Ghana has, at the national level, committed to a One Health approach in recent years (37,70,71), establishing a multi-sectoral Technical Working Group under the National Disaster Management Organization in 2018 to coordinate zoonotic disease efforts (21). Since 2020, further efforts have aimed to formalize and streamline One Health policy implementation. The Rabies Control and Prevention Action Plan – anchored in a One Health framework – has already been launched (72–75), which may explain why rabies-related training in this study was associated with better knowledge and practices, reflecting Ghana’s policy-level commitment to coordinated rabies control. Ghana has also prioritized EVD preparedness following the 2014-2016 West African epidemic (76,77). Although Ghana did not record any Ebola cases, extensive governmental investments in training, infection prevention, and simulation drills likely contributed to the strong Ebola-specific knowledge observed in this study population (76).

However, while intensive outbreak-focused sensitization appears to have effectively embedded knowledge of more familiar threats like EVD and rabies, the findings reveal notable gaps in awareness of emerging zoonoses, alongside limited institutional practices and intersectoral collaboration at the frontline. Though national policies and coordination mechanisms exist in Ghana, their implementation in districts like Ada East appears limited. A recent review echoed this, citing Ghana’s progress is hampered by “weak disease surveillance … and inadequate collaboration across sectors,” underscoring that policies have yet to be fully translated into frontline systems (73). In sum, while Ghana’s One Health policy architecture and outbreak plans offer a strong foundation, a disconnect remains between national strategy and local-level execution.

### Implications for Disease Detection and Surveillance

Knowledge gaps identified in this study carry important implications for early disease detection and surveillance. Limited awareness of common zoonoses increases the risk of misdiagnosis, especially for diseases like influenza or brucellosis that may resemble febrile illnesses such as malaria or typhoid (78). Additionally, poor recognition of endemic zoonoses like salmonellosis and influenza among FHWs in this sample raises concerns that sporadic cases may be missed during initial encounters, as seen in other SSA regions (79,80).

Surveillance systems depend on frontline workers’ ability to identify and report symptoms of zoonotic threats (1,81). Delays in recognition can hinder response and facilitate disease spread, particularly in settings like Ghana, where the risk of spillover events remains high (22,23). Strengthening Ghana’s Integrated Disease Surveillance and Response (IDSR) system will require more than technological or structural upgrades; it also depends on improving FHW knowledge and clinical suspicion of zoonoses (82).

#### Addressing the Knowledge-Attitude Gap

A key finding from this study is the disconnect between FHWs’ knowledge of zoonoses, EVD, and One Health, and their attitudes towards zoonotic threats. While many demonstrated strong factual understanding—particularly of EVD and One Health—93% expressed negative attitudes. This mirrors past epidemics, where knowledge alone failed to overcome stigma, fear, or anxiety (83). Studies from West Africa have shown that even well-informed health workers often held stigmatizing views and exhibited high anxiety during outbreaks (84–86). Such attitudes may impair care delivery. For example, Jalloh et al. documented that during Sierra Leone’s Ebola outbreak, some healthcare providers avoided patients due to fear of infection (87). Similarly, in this study, many supported patient isolation and border closures, and half prioritized personal safety over patient care. These attitudes may reduce willingness to manage suspected zoonotic cases or engage in outbreak response, ultimately delaying containment and reducing care quality.

Findings also point to institutional drivers of fear-based attitudes. Only 29% of FHWs believed their facilities were prepared to manage a zoonotic outbreak. These attitudes were expressed across almost the entire sample; no differences were noted between years of experience, occupational cadre, and facility type, suggesting that these doubts in institutional preparedness and perceived institutional fragility cut across the frontline workforce. Prior research in Ghana supports this finding, citing insufficient PPE, limited training, and unclear risk allowances as contributors to provider reluctance during high-risk events (77,84).

From a behavior change perspective, this finding highlights that effective preparedness must extend beyond knowledge dissemination to foster behavior change and address underlying institutional supports. Without this, even well-informed FHWs may remain hesitant to act in outbreak scenarios.

### Strengths in Individual Practices versus Institutional Collaboration and Training Gaps

This study revealed a clear divide between strong individual-level practices and gaps in intersectoral collaboration and institutional training. Most FHWs reported consistent personal infection control behaviors—68% always used PPE, and 93% routinely notified authorities of suspected zoonotic cases. However, collaborative practices were limited. Only 19% reported routinely engaging with veterinary or environmental health colleagues. This lack of cross-sector interaction risks fragmented responses; for example, treating a zoonotic infection without notifying veterinary authorities may miss opportunities to control the source. These findings suggest that One Health strategies remain under-implemented at the district and community level.

Training gaps were also evident. Fewer than 10% had received training in One Health, and 26% had no formal zoonotic disease training in the past year. Recent literature indicates that many FHWs enter service without exposure to the One Health paradigm or basics of zoonotic disease management, potentially contributing to low preparedness and confidence (41,88). In sum, while individual practices are strong, the broader training infrastructure and intersectoral collaboration may require additional investment.

### Predictors of KAP Outcomes Point to Actionable Levers

The multivariable analyses identified several key predictors of good zoonotic disease knowledge and practices, providing practical leverage points for strengthening frontline capacity against zoonoses in Ada East.

To begin, providers under 35 years had higher odds of good knowledge on zoonoses, EVD, and One Health principles. This trend may reflect recent shifts in educational curricula, where One Health and emerging infectious diseases have gained prominence over the past decade (89,90). Furthermore, this generation of FHWs also came of age during high-visibility outbreaks (e.g., Ebola, COVID-19), likely heightening their awareness. Senior staff (>35 years), who trained before these shifts and may be less engaged with the prominence of e-learning (88,89,91), limiting their exposure to such information and meriting targeted refresher courses to close this generational gap.

Staff who received training delivered by colleagues inside their own facility reported better zoonotic disease prevention practices, demonstrating the value of on-site, facility-specific, and peer-led instruction. Local in-service training sessions may allow frontline teams to troubleshoot in real time, adapt protocols, and tailor practices to local realities— an effect echoed in a Ghanaian study showing that capacity building via in-service training boosts both confidence and competence in infectious disease response (88). Designating in-house “One Health champions” to mentor peers through regular case discussions may prove beneficial. By contrast, trainings offered by governmental agencies were not linked to good knowledge in this data, suggesting that “top-down” approaches in training may not readily translate into better knowledge given the diversity of cases experienced by FHWs in Ada East (82).

Prior rabies training was shown to predict good zoonotic disease knowledge and preventative practices, potentially indicating that existing rabies-specific training translates well into awareness and usable skills when managing potential zoonotic disease cases in Ada East (73,74). Rabies remains a familiar, high-stakes threat in Ghana, with 112 human rabies deaths per year (92). Expanding existing rabies training to include modules on other locally endemic zoonoses and One Health concepts could deliver efficient, disease-agnostic capacity building and dispel lingering stigma that knowledge alone cannot.

Providers who were exposed to public sources of climate change information and frequently engaged with climate change content demonstrated better zoonotic knowledge and practices. Linking environmental shifts to spill-over risk might broaden preventative thinking (93,94). Integrating climate-health discussions into routine zoonotic disease education programs may, therefore, sensitize providers to broader disease ecology and behavior change.

### Limitations and Future Research Directions

This study offers a granular view of frontline readiness using a structured KAP instrument spanning multiple zoonotic disease domains. By surveying FHWs from 18 facilities on different days and shifts, we increased the generalizability of this study to reflect multiple workplace environments in Ada East. Defined KAP cut-offs strengthened the comparability and internal validity of this data. However, our recruitment strategy may have preferentially included FHWs who were available and willing during site visits, introducing selection bias. To mitigate this, visits occurred at various hours and on multiple days to reach different staff schedules. Self-reported practices remain vulnerable to social-desirability bias. The cross-sectional design limits causal inference. Future longitudinal or cohort studies should examine FHWs’ understanding of how climate change influences zoonotic disease risk to better assess whether integrated One Health training leads to sustained improvements in knowledge and practices.

## Conclusion

FHWs in Ada East know a great deal about Ebola and engage in personal protective measures, yet they remain fearful of and under-exposed to the broader spectrum of zoonotic threats facing Ghana. Bridging this knowledge-attitude-practice gap demands more than knowledge dissemination: it requires One Health teams, training, and educational programs on locally relevant zoonoses, and risk-communication strategies that boost confidence while dismantling stigma. Strengthening these competencies will improve case finding and reporting, enabling Ghana’s surveillance system to detect emerging zoonoses before they spiral into larger outbreaks.

## Data Availability

Data are only available upon reasonable request and data transfer requires a written agreement approved by the Ghana Health Service and the Pennsylvania State University. Data inquiries can be sent to Abebayehu N. Yilma at.

## Acknowledgements

The authors would like to thank all the frontline healthcare workers who generously shared their time and insights, as well as the Ada East district health facility leadership for their support during data collection. We are also thankful to the contributions of the local research assistants and field staff involved in data entry and community engagement. It is our hope that these findings inform meaningful improvements in health system preparedness and response in Ada East. This research has been accepted for poster presentation at the American Public Health Association (APHA) 2025 Annual Meeting; however, no abstract has been formally published at the time of manuscript submission.

